# An AI-assisted Online Tool for Cognitive Impairment Detection Using Images from the Clock Drawing Test

**DOI:** 10.1101/2021.03.06.21253047

**Authors:** Samad Amini, Lifu Zhang, Boran Hao, Aman Gupta, Mengting Song, Cody Karjadi, Honghuang Lin, Vijaya B. Kolachalama, Rhoda Au, Ioannis Ch. Paschalidis

**Author notes:** Corresponding author: Ioannis Ch. Paschalidis, 8 St. Mary’s St Boston, MA 02215.

## Abstract

**Background:** Widespread early dementia detection could drastically increase clinical trial candidates and enable early interventions. Since the Clock Drawing Test (CDT) can be potentially used for diagnosing dementia related diseases, it can be leveraged to devise a computer-aided screening tool.

**Objective:** This work aims to develop an online screening tool by leveraging Artificial Intelligence and the CDT.

**Methods:** Images of an analog clock drawn by 3, 263 cognitively intact and 160 impaired subjects were used. First, we processed the images from the CDT by a deep learning algorithm to obtain dementia scores. Then, individuals were classified as belonging to either category by combining CDT image scores with the participant’s age.

**Results:** We have evaluated the performance of the developed models by applying 5-fold cross validation on 20% of the dataset. The deep learning model generates dementia scores for the CDT images with an Area Under the ROC Curve (AUC) of 81.3% *±* 4.3%. A composite logistic regression model using age and the generated dementia scores, yielded an average AUC and average weighted F1 score of 92% *±* 0.8% and 94.4% *±* 0.7%, respectively.

**Discussion:** CDT images were subjected to distortion consistent with an image drawn on paper and photographed by a cell phone. The model offers a cost-effective and easily deployable mechanism for detecting cognitive impairment online, without the need to visit a clinic.

## 1. Introduction

As the world population ages, cognitive decline is becoming a prevalent condition. Cognitive decline is present in several neurodegenerative disorders such as *Alzheimer’s Disease (AD)*, vascular dementia, and Parkinson’s disease [1]. In the U.S., (*i*) the cost associated with *AD and Related Dementias (ADRD)* has been estimated to be $305 billion in 2020, expected to rise to as much as $1.1 trillion by 2050, and (*ii*) more than 5 million individuals are living with AD, with AD deaths increasing by 146% between 2000 and 2018 [2]. Worldwide, it is estimated that more than 50 million are living with dementia [3].

The standard approach to evaluate the severity of cognitive decline of an individual includes *Neuro-Psychological (NP)* exams, which have traditionally been conducted via in-person interviews to measure memory, thinking, language, and motor function. However, this approach can be expensive, time-consuming, and limited in availability to subjects with lower income and/or belonging to a racial or other underrepresented minority. With the ongoing COVID-19 pandemic, access to medical facilities for non life-threatening conditions has been curtailed and medical care has shifted to virtual, online visits. Such an approach is highly desirable for dementia screening. In addition to broader and more equitable access to care, it can lead to earlier diagnosis and drastically increase the pool of candidates for ADRD clinical trials, possibly accelerating the search for effective treatments.

With this motivation, this work develops an AI-assisted approach capable of detecting cognitive decline based on the venerable *Clock Drawing Test (CDT)*. In this test, subjects are asked to draw the face of an analog clock showing ten minutes past eleven. CDT is considered robust against cultural biases and language and provides insight into the mechanisms underlying cognitive dysfunction, including comprehension memory, numerical knowledge, and visuoperceptual skills [4, 5]. Given the sensitivity of CDT in cognitive screening, numerous attempts have been made to exploit the full potential of CDT in identifying dementia. For instance, the sensitivity of the CDT was investigated in [1] to monitor and distinguish the evolution of cognitive decline in different cognitive domains. In [6], the authors found CDT useful in cognitive impairment screening using the fact that the CDT score correlates with the severity of global cognitive impairment, as assessed by the *Mini-Mental State Examination (MMSE)* score and the Hasegawa dementia scale. A low CDT score was also associated with progression to dementia, with the association being independent of the MMSE score [7, 8].

Options such as *digital Clock Drawing Test (dCDT)* technology, where the drawing is traced by a digital pen, enable the examination of a detailed neurocognitive behavior as it unfolds in real-time; a capability that cannot be obtained using a traditional pen and paper [9, 10]. Recently, researchers have been trying to utilize machine learning methods for early prediction of ADRD. By using a dCDT, a machine learning approach based on non-interpretable boosted decision trees was able to outperform scoring systems used by clinicians [11], reaching an Area Under the Receiver Operating Characteristic (ROC) Curve (AUC) of 93% using the entire battery of features provided by dCDT. The AUC drops to 83% for simpler, interpretable models. [12] introduced a classification task to classify mild cognitive impairment subtypes and AD using 350 dCTD features, with accuracies ranging from 83% to 91%. Others have leveraged medical imaging, which is expensive and requires an in-person visit to an imaging facility. Deep learning was applied to predict progression to AD based on hippocampal *Magnetic Resonance Imaging (MRI)* and other baseline clinical features [13], achieving an AUC of 86%. Deep learning on brain MRI images was also used in [14], which, when combined with non-imaging features, produced F1 scores ranging from 63.3% to 96.5%. Furthermore, a robust deep learning classifier was adopted in [15] to identify the different stages of mild cognitive impairment based on MRI and *Positron Emission Tomography (PET)* [16, 17], with accuracies ranging from 57% to 91%. In general, and as we elaborate on later, accuracies are not the most appropriate metric for assessing performance since AD/ADRD datasets are heavily imbalanced (only a small fraction of subjects have dementia).

The above studies rely on a large collection of features available through NP tests, dCDT, blood biomarkers (e.g., apolipoprotein genes), or medical imaging, thus requiring expensive resources and in-person visits. These technologies, even the digital pen, make the cost prohibitive for low-resourced health care environments and perpetuate persistent health disparities in global testing. In the context of using features from these tests for AI-assisted detection, they also embed inherent biases, further exacerbating the widening gap across global populations in health care knowledge and practice. The proposed approach uses only photos of CDT and the age of the participant, offering an inexpensive and remote screening technique that can be made available online. To that end, our method processes CDT images through a deep *Convolutional Neural Network (CNN)* [18, 19] classifier and combines the output scores with age in an ensemble, logistic regression-based classifier.

## 2. Materials and Methods

### 2.1. Clinical setting and data sources

The data used in this study were collected by the *Framingham Heart Study (FHS)*, the longest ongoing longitudinal study of chronic disease [20]. Since 2011, the FHS has adopted digital pen technology to capture pen and paper NP tests, including the CDT. In the FHS dataset, two different clock images are collected: (*i*) one where the subjects are told to draw an analog clock showing ten minutes past eleven (*command clock*), and (*ii*) one where they are asked to copy the image of such a clock shown to them (*copy clock*). Additional information available includes sex, age, race, and presence of Apolipoprotein E (ApoE) genes. All subjects were evaluated by trained examiners. For those subjects identified as showing symptoms of cognitive decline, dementia diagnosis was reached by consensus of at least one neurologist and one neuropsychologist; the dementia surveillance and diagnostic procedures have been previously outlined in [5, 21]. The entire dataset for all participants was anonymized prior to analysis. All participants have provided written informed consent and study protocols and consent forms were approved by the Boston University Medical Campus Institutional Review Board.

### 2.2. Data preparation

The original dataset contains information about 3, 423 participants. The dataset attributes consist of participant demographic data, ApoE gene information, command and copy clock drawings, as well as the dementia diagnosis. The clock drawings are stored in *.csk* format as they are recorded using the digital pen [22]. Therefore, a pre-processing pipeline was created to convert the *.csk* files into the clock images of size (128, 128, 3), which are three-channel images with 128 *×* 128 pixels. To normalize the data, the value of each pixel was divided by 255 to rescale pixel values into the [0, 1] range. Furthermore, we conducted data augmentation on the original images by randomly applying *±*10 degrees rotation, *±*15 percent zoom, *±*10 percent width and height shift, and *±*10 percent shear. Data augmentation enables us to develop a deep learning model robust against image distortions consistent with someone drawing the clock using pen and paper and taking a digital photograph (e.g., using a cell phone). By disproportionally augmenting the non-dominant class of images, we also mitigate class imbalance – 95.3% of participants have no cognitive impairment – enabling training of the deep learning model in a balanced fashion without under-sampling the dominant class.

### 2.3. Statistical analysis

The composition of the dataset along with basic statistics is reported in Table 1. The 2nd column provides information on participants who were labeled as normal and the 3rd column corresponds to participants diagnosed as cognitive impaired (Cognitive Impairment, No Dementia – CIND), or with clinical dementia (mild, moderate, severe). We report self-reported gender, age statistics (mean *±* standard deviation for each cohort), race, dementia diagnosis severity, and the type of ApoE (E2/E3/E4) genes for both copies of the gene. In the 4th column we report the *p*-value for each variable associated with the null hypothesis that the two cohorts have the same distribution of the variable. Hence, a low *p*-value implies that the distribution of the feature is different in each cohort, leading us to reject the null hypothesis. For age, we employed the Kolmogorov-Smirnov (K-S) test [23] whereas we used the Chi-square test for the categorical features [24]. It can be seen that only age shows significant difference among the two cohorts, leading us to use only age and CDT images in the proposed ensemble model. As we will see, adding additional variables leads to minimal improvement. An additional advantage of using only CDT images and age is that these features are easily obtained remotely without the need to visit a clinic.

**Table 1:** Summary of the variables in the FHS dataset. Diagnosis scores 0, 0.5, 1–1.5, 2–2.5, and 3 are defined as normal cognition, CIND, mild dementia, moderate dementia, and severe dementia, respectively.

| Variable | Assessment |  | *p*-value |
| --- | --- | --- | --- |
|  | Normal<br>(n=3263) | CIND/Dementia<br>(n=160) |  |
| Diagnosis |  |  | – |
| 0 | 3263 (100%) | 0 (0%) |  |
| 0.5 | 0 (0%) | 96 (60%) |  |
| 1–1.5 | 0 (0%) | 38 (24%) |  |
| 2–2.5 | 0 (0%) | 24 (15%) |  |
| 3 | 0 (0%) | 2 (1%) |  |
| Age | 61.8 ± 13.2 | 82.1 ± 7.3 | <0.0001 |
| Gender |  |  | 0.95 |
| Female | 1773 (54.3%) | 86 (53.75%) |  |
| Male | 1490 (45.7%) | 74 (46.25%) |  |
| ApoE |  |  | 0.12 |
| 22 | 19 (0.6%) | 1 (0.6%) |  |
| 23 | 399 (12.2%) | 19 (11.9%) |  |
| 24 | 63 (1.9%) | 6 (3.8%) |  |
| 33 | 2011 (61.6%) | 87 (54.4%) |  |
| 34 | 593 (18.2%) | 40 (25%) |  |
| 44 | 45 (1.4%) | 4 (2.5%) |  |
| Race |  |  | 0.24 |
| Asian | 86 (2.6%) | 1 (0.6%) |  |
| Black | 87 (2.7%) | 1 (0.6%) |  |
| Hispanic | 80 (2.5%) | 2 (1.2%) |  |
| White | 2957 (90.6%) | 156 (97.5%) |  |
| others | 53 (1.6%) | 0 (0%) |  |

### 2.4. Development of the model

We formulated the cognitive impairment detection system as a classification task in which the model seeks to make a binary decision about whether the participant is cognitively impaired – dementia diagnosis score above zero. Since there exist insufficient data for subjects with either CIND or dementia, a single class is representing CIND/dementia in our proposed model.

Given that CNN deep learning models, which include many hidden layers and millions of parameters, require a large number of images to be trained, we adopted a transfer learning approach starting from a pre-trained CNN. Transfer learning is widely used in medical image analysis and natural language processing applications [25, 26]. As the backbone network of our proposed method, we selected the lightweight MobileNet V2, which can be trained fast and is very suitable for embedded devices [27, 28]. We modified the MobileNet V2 model using the CDT images in the training set. To that end, we detached the *fully-connected* layer and attached a global average pooling operation to convert the feature map into a smaller size by taking the average value from the spatial dimension of the feature map. Global average pooling avoids overfitting and provides a more robust model against spatial translations [29]. A *softmax* layer is attached to the deep learning model to predict the probability distribution of each class. In the training procedure, all the layers of the MobileNet V2 trained on the ImageNet [30] were frozen, except the softmax layer. Since the MobileNet V2 is based on a three-channel CNN, we used command CDT images of size (128, 128, 3) to train the modified deep learning model. ^1^ The participants tend to make more mistakes in the command clock drawing task compared with the copying task (cf. Section 4), hence, the command clock images can reveal more types of image defects associated with cognitive impairments.

Once the training process of the deep learning model is completed, the scores corresponding to test command and copy images can be generated by feeding them into the model. Finally, image scores and the age of every participant were used to train a logistic regression model to make predictions; a schematic of the approach is shown in Figure 1. The entire model was implemented using the python deep-learning *Keras* library with a *Tensorflow* backend.

**Figure 1:**
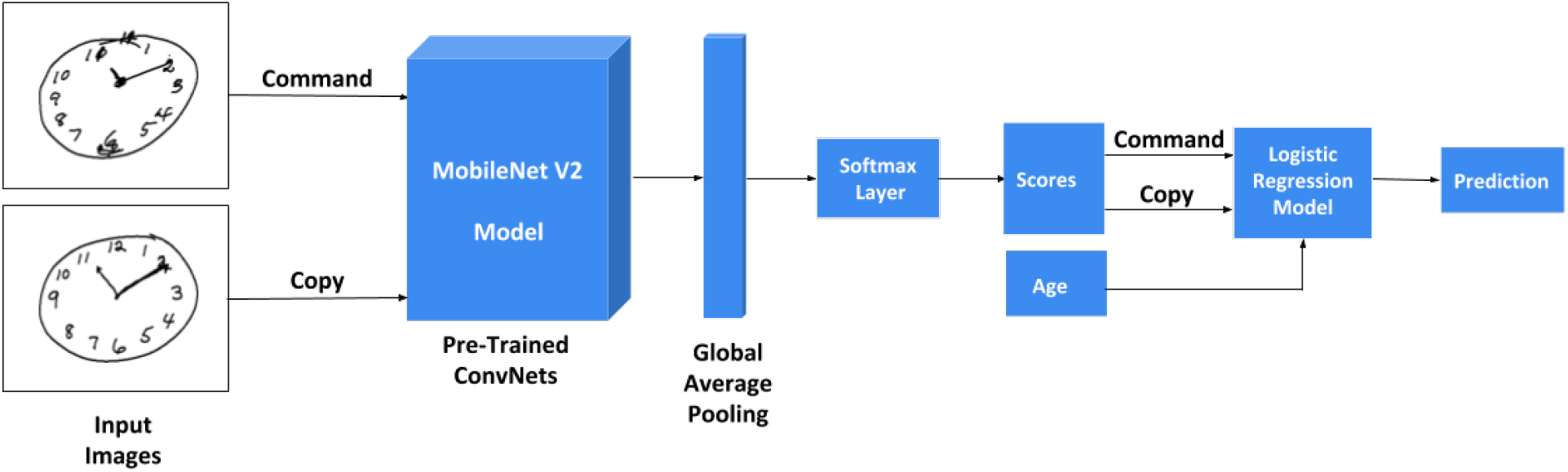
Online screening for cognitive impairment.

### 2.5. Validation and performance metrics

Data were randomly split into 5 folds using stratified *k*-fold cross validation. Specifically, the models were trained on the 4 folds and tested on the 5th – test – fold. The process was repeated five times, each time with a different fold retained for testing, and the average and standard deviation (std) of all performance metrics on the test set over these five runs was obtained.

Performance metrics included the *Area Under the Receiver Operating Characteristic (ROC) Curve (AUC)* and the *weighted F1 score* [31]. The ROC plots the True Positive Rate (TPR, a.k.a. recall or sensitivity) against the False Positive Rate (FPR, equal to 1 minus the specificity). The F1 score is the harmonic mean of precision and recall. Precision (or positive predictive value) is defined as the ratio of true positives over true and false positives. The F1 score is calculated by

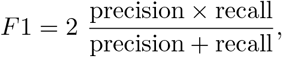

and the closer it is to 1 the stronger the classification model. In this work we report the *weighted F1 score*, which is computed by weighting the F1-score of each class by the number of participants in that class.

## 3. Results

As detailed in Section 2.5, we trained and validated the proposed classifier using 5-fold cross-validation. The results are summarized in Table 2. We report ROC AUC, weighted F1 score (W-F1), and accuracy (Acc), the latter mainly for comparison purposes with results reported in the literature and surveyed in Section 1. We note however, that accuracy in binary classification with a highly imbalanced dataset is not an appropriate metric, since predicting all subjects as being normal would lead to a high accuracy.

**Table 2:** Results on the test set (mean *±* std over the five runs).

| Methods | AUC % | W-F1 % | Acc % |
| --- | --- | --- | --- |
| **Cmnd CDT** | 81.3 ± 4.3 | 92.8 ± 1.0 | 87.8 ± 1.2 |
| **Ensemble** | 92.0 ± 0.8 | 94.4 ± 0.7 | 95.1 ± 0.3 |
| **Full** | 92.3 ± 0.8 | 94.4 ± 0.5 | 95.4 ± 0.5 |
| **Full\ApoE** | 91.9 ± 0.8 | 94.6 ± 0.7 | 95.1 ± 0.4 |
| **Age, Cmnd CDT** | 91.8 ± 0.7 | 94.3 ± 0.7 | 95.1 ± 0.4 |
| **Age** | 89.3 ± 1.2 | 93.4 ± 0.6 | 95.1 ± 0.4 |

In the first row of Table 2, we report the performance of the classifier that uses just the command CDT (Cmnd CDT) image of a subject. The second row reports the performance of our proposed *ensemble model* obtained by using logistic regression with features the deep learning scores of command and copy CDT images and the subject’s age. The third row, corresponds to the full model, which uses command and copy CDT images, age, gender, ApoE, and race. The fourth row corresponds to a model using all these features, except ApoE. The fifth column reports the performance of a model using only age and the command CDT image. Finally, the last row, reports the performance of a model based just on age. In all these models, gender, ApoE, and race features, were encoded using one-hot encoding, i.e., creating a binary variable for each category. Figure 2 plots the ROC curves for the two models with the highest average AUC (full and ensemble), and, for comparison purposes, the corresponding curve for the age-based model.

**Figure 2:**
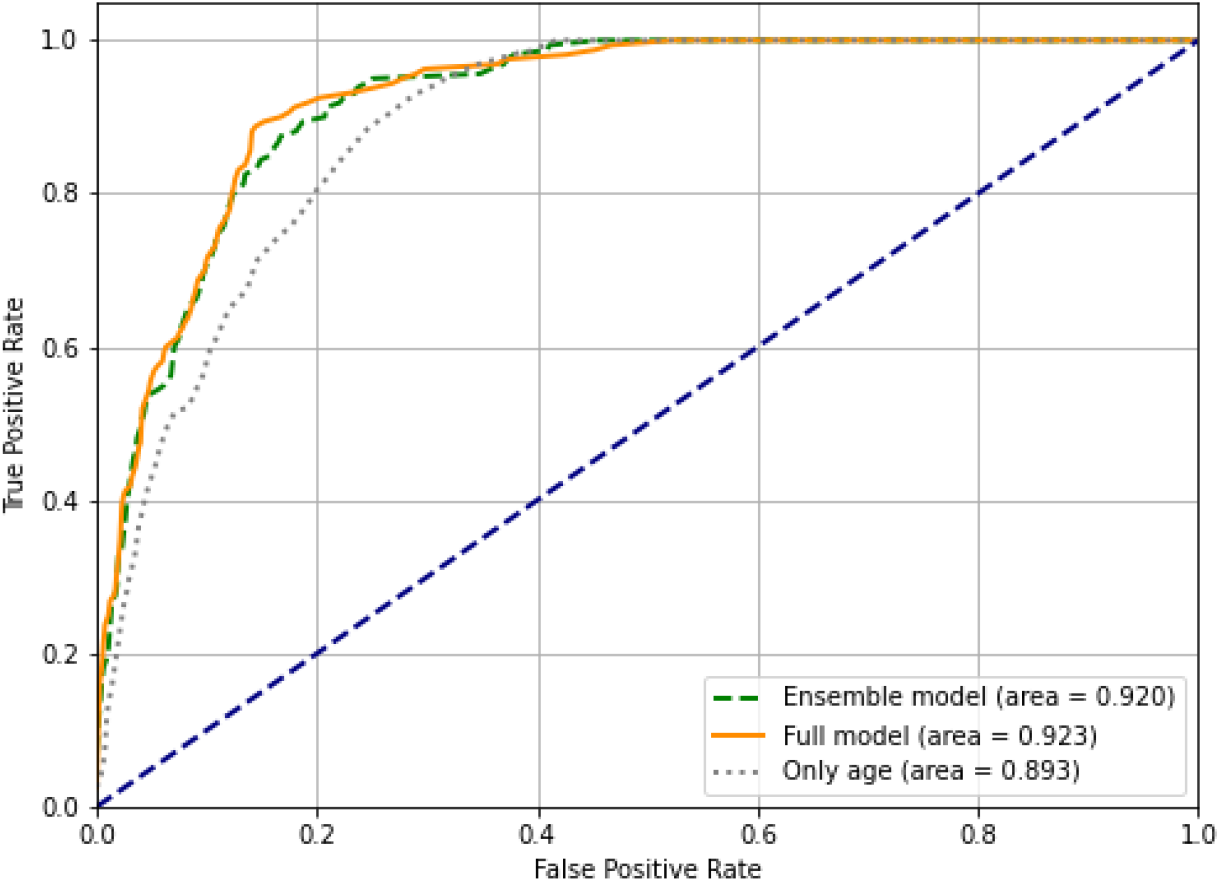
ROC curves for three models: the proposed ensemble model, the full model, and the model based only on age. We plot the average TPR and FPR over the five folds.

## 4. Discussion

The proposed *ensemble* model can be employed to offer virtual cognitive impairment screening using only CDT images drawn on pen and paper, and captured via a cell phone, and the age of the subject. The out-of-sample AUC and weighted F1 scores we report indicate strong predictive power, at least within the context of related metrics from other methods we outlined in Section 1.

An important aspect of the method we used for training the deep learning models was *transfer learning*. We started from a deep learning image classifier previously trained on a large number of generic images, which, apparently, has the ability to identify useful features of presented images. Thus, with limited training using our CDT images, the deep learning model adapted quickly to score CDT images and produced scores representing the likelihood of the subject being cognitively impaired. Just using command CDT images yields a classifier of moderate strength (cf. first row of Table 2, AUC of 81.3%, on average). Combining command and copy CDT images with age using logistic regression yields a strong screening tool with an AUC of 92%, on average, and a weighted F1 score of 94.4%, on average.

Figure 3 reports the coefficients of the three features used by the logistic regression ensemble model. The coefficients are comparable as the scores of the CDT images and age were normalized by subtracting the mean and dividing by the standard deviation. It can be seen that age and the command CDT image are contributing more to the decision than the copy CDT image.

**Figure 3:**
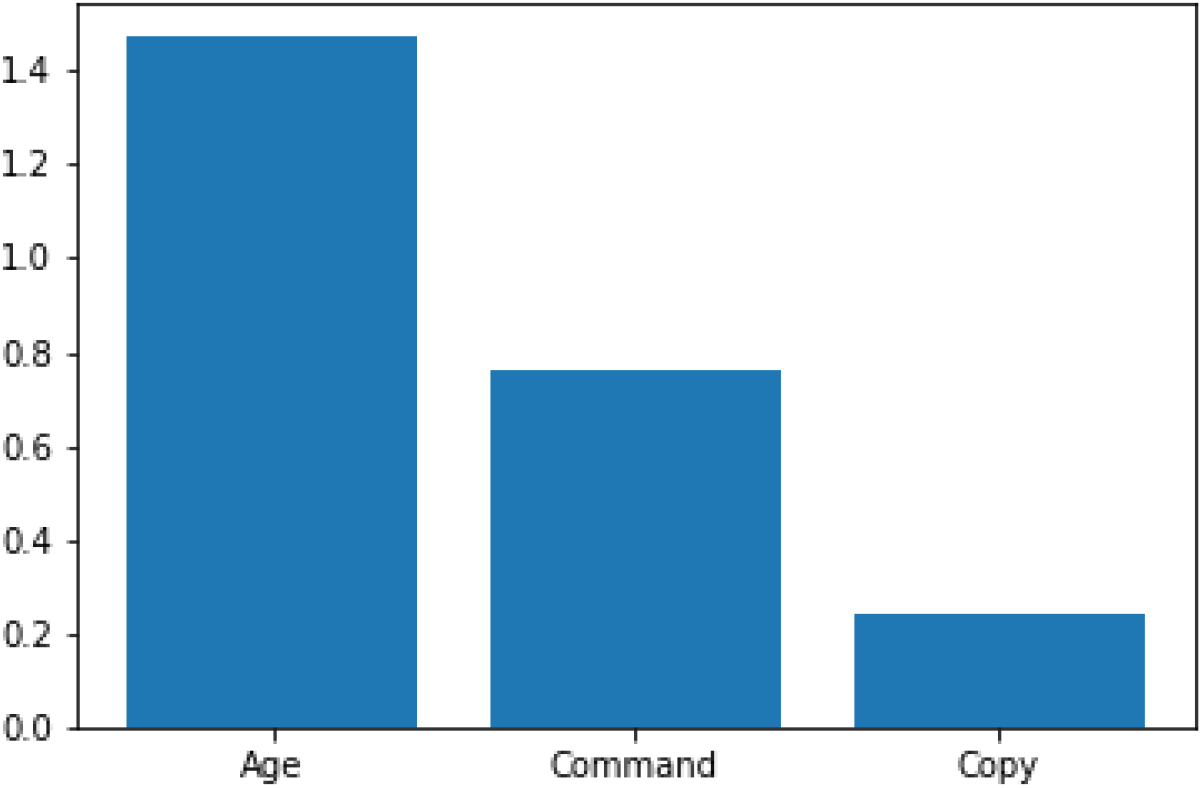
Logistic regression coefficients, indicating the relative predictive importance of the three features.

The ensemble model performs closely to the full model (cf. Table 2) which uses all features. However, the full model is not amenable to online screening as ApoE genotyping requires laboratory testing (typically, a blood sample). From Figure 2, it can also be seen that the ROC curves of the full model and the ensemble model essentially coincide for low values of the FPR (below 15%), that is, within the range one may want to operate. Interestingly, adding to the ensemble model gender and race (i.e., full model without ApoE), leads to worse performance (AUC of 91.9% vs. 92%, on average), confirming the low discriminatory power of these additional features, which was also suggested by their *p*-values listed in Table 1. Using just age and command CDT images performs slightly worse than the ensemble model, confirming the findings of Figure 3. Table 2 also indicates that a model based just on age performs relatively well; average AUC of 89.3% vs. 92% for the ensemble model, yielding a difference of 2.7%. This is consistent with related findings in [14] where a model based on age, gender, and MMSE had an average F1-score 1.4% lower on their internal validation dataset than the fusion model which also used a brain MRI. It is useful to compare the average TPR (sensitivity) of the ensemble model, the full model, and the age model for low values of the average FPR (high specificity). This comparison is shown in Table 3. For instance, at 10% FPR, the TPR of the ensemble model is 11.8% higher compared to the age model. Putting this difference in context, suppose we were interested in screening for a nationwide clinical trial all 5 million or so individuals in the U.S. estimated to be suffering from Alzheimer’s [2]. Setting FPR to 10%, about 3.57 million would qualify with the ensemble model vs. 2.98 million with the age model, missing a non-trivial number of about 600,000 subjects with the latter. A similar perspective is gained by considering how many individuals one should screen to assemble a clinical trial with about 1600 subjects (similar in size to the EMERGE aducanumab trial by Biogen [32]). Using an FPR of 5%, and assuming the CIND and dementia incidence rate is 4.7% as in our dataset, it follows that one needs to screen 78,439 people with the age model compared to 62,578 with the ensemble model, namely, 15,861 less. Clearly, these differences imply significant differences in cost.

**Table 3:**
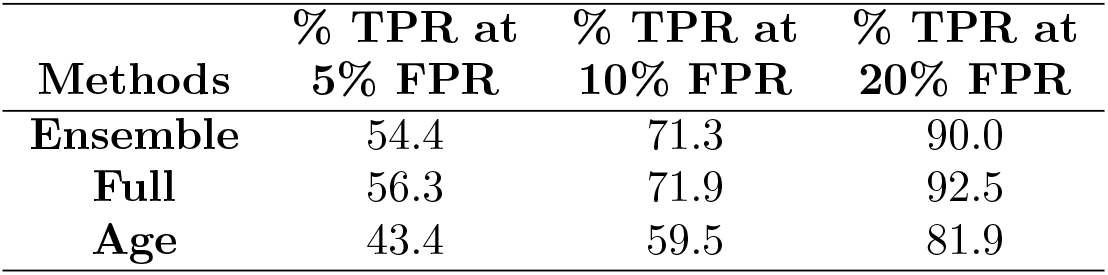
Average True Positive Rate (TPR, or sensitivity) for average False Positive Rates (FPR) at 5%, 10%, and 20%.

| **Methods** | **% TPR at**<br>**5% FPR** | **% TPR at**<br>**10% FPR** | **% TPR at**<br>**20% FPR** |
| --- | --- | --- | --- |
| **Ensemble** | 54.4 | 71.3 | 90.0 |
| **Full** | 56.3 | 71.9 | 92.5 |
| **Age** | 43.4 | 59.5 | 81.9 |

In the present study, the clock drawing images were collected from the FHS using a digital pen. The size of the images used for training was reduced to 128 *×* 128 pixels. Furthermore, the data augmentation described in Section 2.2 empowers the deep learning features to become robust to various forms of image distortion that could be introduced by drawing the images using pen and paper and capturing them using a cell phone.

A limitation of the study is that we do not have access to actual cell phone-captured images, which would provide the ultimate test for the proposed screening approach. An additional limitation is that the FHS does not comprehensively conduct dementia review of all participants; thus, it is possible that some subjects classified as cognitive normal are in the early CIND stages. We submit that this limitation would lead to a bias toward the normal class and may reduce the possibility of a false positive diagnosis.

### Diagnostic Tool Availability

We have made our code publicly available. ^2^ Upon acceptance of the paper and before publication, we plan to make the proposed cognitive impairment assessment tool available to the community online. Specifically, we will set up a web site where anyone could submit two clock images (command and copy) and age and receive the corresponding probability of cognitive impairment.

## Data Availability

The dataset is coming from Framingham heart study and is not available to public.

## Acknowledgments

The research was partially supported by the NSF under grants DMS-1664644, CNS-1645681, and IIS-1914792, by the ONR under grant N00014-19-1-2571, by the NIH under grants R01 GM135930 and UL54 TR004130, by the DOE under grant DE-AR-0001282, by the Framingham Heart Study’s National Heart, Lung, and Blood Institute contract (N01-HC-25195; HHSN268201500001I), by the NIH National Institute on Aging (AG008122, AG016495, AG033040, AG054156, AG049810, AG062109), by the Alzheimer’s Association under grant AARG-NTF-20-643020, and by Pfizer. Rhoda Au is a scientific advisor to Signant Health and consultant to Biogen. There is no declaration from other authors.

## Footnotes

1 The input size of the MobileNet V2 can be adjusted as needed.

2 https://github.com/noc-lab/CDT.

